# From Evidence to Influence: Media’s Role in Informing Urban Health Policies in Bangladesh and Nepal

**DOI:** 10.64898/2026.10.14.26365501

**Authors:** Sabrina Mustabin Jaigirdar, Sulata Karki, Shreeman Sharma, Rumpa Akter, Sabina Marasini, Rumana Huque, Sushil Baral, Helen Elsey, Zahidul Quayyum

## Abstract

**Background:** Urban inequities are often masked by the misleading notion of an urban advantage, despite persistent unequal access to quality healthcare services. Moreover, negative perceptions about the urban poor and slum-dwellers persist, despite their substantial role in driving and holding up the urban economy. It is thus essential to work with the media to bring visibility to these realities, raise public awareness, and inform more equitable health policies. In low- and middle-income countries (LMICs), evidence remains scarce on how health-related information from media outlets influences policy decisions. This study aimed to understand how journalists get information, engage with stakeholders in the health systems, and contribute to influencing urban health discourse in Bangladesh and Nepal.

**Methods:** We conducted a cross-country exploratory qualitative study comprising 30 key informant interviews from Bangladesh and Nepal, including 17 journalists, 10 health policymakers, and 3 public health advocates. Participants were purposively selected to ensure diversity across media types, gender, and professional roles. Interviews were conducted between September 2024 and March 2025. Data were transcribed, translated, and analyzed thematically using NVivo software. Cross-country comparative analysis was carried out to identify common themes and contextual differences.

**Findings:** Across both countries, participants relied on multiple and diverse information sources. Government data were often prioritized as a primary source of information, although they were shaped by political influences and accessibility constraints. Research evidence was valued but underutilized due to difficulties in interpreting scientific articles. Urban health issues received limited priority in health news reporting due to journalists’ constraints in covering multiple issues and financial constraints for in-depth coverage. Additional challenges included inadequate health and urban health-related training, limited fellowship opportunities for investigative reporting, and restricted access to data, while health policymakers and public health advocates frequently questioned the credibility and use of media-reported information. Media engagement in policymaking largely remained indirect and issue-focused or event-driven. However, consistent media coverage raised public awareness and generated public pressure and, at times, indirectly influenced health policy decisions such as immediate inspection after reporting on pesticides, installation of oxygen plants during the pandemic, dengue surveillance, and tobacco control measures and actions.

**Conclusion:** In Bangladesh and Nepal, news media play a significant but indirect role in shaping urban health policy decisions. Enhancing journalists’ capacity, improving access to credible information and data, and continuous engagement of stakeholders could strengthen evidence-informed urban health policymaking. Furthermore, establishing greater independence of journalists and government and community recognition of their responsible health reporting could help and motivate the media to contribute more meaningfully to public debate and policy reform processes.

## Introduction

Unplanned rapid urbanization, particularly in low- and middle-income countries (LMICs), over the years has reshaped urban environments and contributed to a growing range of public health challenges. In many cities, rapid and often unplanned urban growth has contributed to worsened living conditions, increased mobility pressures, environmental health hazards, and unsafe communities [1, 2]. In addition, urban residents face a triple burden due to the rising noncommunicable diseases (NCDs), infectious disease outbreaks, and increased risk of violence and injuries [3]. These overlapping challenges limit policymakers’ ability to adapt to evolving contexts and respond effectively through policy interventions. In countries such as Bangladesh and Nepal, urbanization has led to lifestyle changes and living environments that place additional pressure on already constrained urban health systems [2, 4]. Furthermore, a pronounced equity gap exists in the urban setting, as wealthier populations benefit from better access to healthcare and services, while marginalized groups often face worse health outcomes than rural residents [2], relying on a fragmented and informal urban health system, such as local pharmacies [5]. While public healthcare is meant to be free, patients still pay for medicines, tests, and other hidden costs [6]. Addressing such complex urban health challenges requires multisectoral approaches that extend beyond reforms within the health sector and tackle broader social determinants of health [2]. Among these sectors, the news media play a critical role by raising public awareness and knowledge, strengthening stakeholders’ communication, helping evidence translate into public discourse, and influencing public health policy agendas [7–9]. Media are particularly important in urban contexts, where there are persistent misconceptions surrounding the urban advantage, including the assumption that all urban residents have equal access to quality healthcare despite major inequities. In reality, marginalized urban populations often experience poor living conditions and barriers to accessing healthcare and other services [10]. By shaping public discourse and drawing attention to neglected issues, the media can play an important role in reducing urban health inequities and increasing accountability. Evidence shows media reporting positively influenced health outcomes by shaping public attention, framing risk, and exerting pressure on policymakers, which was evident during COVID-19 [11, 12]. A systematic review found that media interventions, such as news coverage and campaigns, raised policymakers’ awareness and supported policy adoption or enforcement [13]. Scholars argue that media concentration in cities strengthens journalism’s influence in policy debates by setting agendas and pressuring governments to respond [14].

The media landscapes in both countries are evolving, particularly with the growth of digital platforms, and the role of the media in providing accurate health information is increasingly important [15]. In Bangladesh, health journalism has experienced significant progress in recent years, supported by the rapid growth of print, television, and online media, which has expanded the reach of health information and public awareness [15, 16]. Similarly, health journalism in Nepal has shown progress, and journalists have increasingly covered health topics over the past five years [17]. At the same time, digital and social media have become a powerful means to influence the opinions of young people and their participation, which was manifested in recent youth-led political movements in both countries [18, 19].

In both countries, national urban health strategies or policies have been introduced to address the growing needs of urban dwellers; many urban poor remain excluded from health services due to unstable housing, informal employment, and their absence in government records, as many are not formally registered within official systems [6, 20–25]. Mass media have the potential to complement the gaps while fulfilling the role of the press in promoting the voice of these disadvantaged groups in the policymaking process and strengthening the accountability of governments to citizens [26]. Urban health issues, especially those affecting the marginalized communities need more attention in both the media and policy discussions, as these communities, especially people living in informal settlements, are often left out of health data, giving an incomplete picture of the city’s health [27]. However, journalists faced challenges in health news reporting. Studies show that health reporting in news media often contains information that is not backed up by sufficient evidence and scientific reasoning. This is partly due to the lack of necessary skills and knowledge among journalists and media professionals [16, 17]. Available training is also not adequate, especially in understanding and interpreting health narratives, and there’s a lack of technical skills in understanding research methods needed for reporting based on evidence and data [16, 28]. Despite these limitations, exposure to news media has still been linked to policy reforms that have led to positive public health outcomes in both high-income and LMIC settings [13, 29]. Evidence from South Asia, such as Bangladesh, India, Nepal, and Pakistan, shows that women with access to television, radio, or newspapers were significantly more likely to receive antenatal care, give birth with skilled personnel, and access postnatal care [30]. These findings highlight the dual role of media in both informing the public and shaping policy priorities by influencing public discourse and sensitizing key stakeholders [13]. In this context, strong and constructive relationships between journalists and health policymakers can improve journalists’ access to credible, evidence-based sources of information, particularly for health reporting [31].

While existing literature acknowledges the role of media and research in knowledge dissemination, public awareness and stakeholder communication, limited evidence exists on how news media integrate scientific and experiential knowledge to influence urban health policy discourse [13, 14]. Moreover, a persistent gap remains between evidence-informed policy decisions and their translation into action [32]. The media can play an important oversight role by promoting good governance and accountability through the provision of accurate, balanced, and timely information [33]. However, despite this recognized potential, there remains a limited body of research in LMICs examining the role of media in evidence translation [34] in the context of urban health. Additionally, gaps also remain in understanding stakeholder dynamics, particularly how journalists, public health advocates, and health policymakers engage with each other in urban health policy formulation processes and evidence dissemination. Therefore, this study aimed to explore the role of news media in shaping the urban health agenda and policy reforms in Bangladesh and Nepal. Specifically, it examined journalists’ urban health reporting practices and challenges, policymakers’ use of media-reported information, and the engagement of media and policymakers in shaping health policy decisions.

## Methods

### Study design, setting, and sample

This study employed an exploratory qualitative approach. A total of 30 key informant interviews (KIIs) were conducted in Bangladesh and Nepal, involving 17 journalists, 10 policymakers from the health sector, and 3 public health advocates (individuals promoting and advocating for policies and actions to improve population health and address health inequities).

The journalists were from various news media outlets, primarily engaged in health reporting and other related beats. The health policymakers from Bangladesh included representatives from the health department under the Ministry of Local Government, the Ministry of Health and Family Welfare, the Dhaka North City Corporation, and the Bangladesh Institute of Planners. Public health advocates in Bangladesh and policymakers from Nepal were representatives from academia, non-governmental and government organizations, and federal-level public hospitals.

The selection of journalists was guided by criteria such as media type (print newspapers, online news portals, and broadcast media); geographic diversity; gender; newspaper popularity; type of ownership, such as public and private; circulation; and language (Bangla, Nepali, and English). Selection of the participants was also guided by mentors and experts. Priority was given to individuals actively engaged in urban health, health planning, policy formulation, advocacy, and decision-making. Participants were selected through purposive sampling, and the principle of saturation was applied to determine the final number of interviews based on the sufficiency of information [35].

### Data collection

A structured topic guide, similar for both countries but contextualized to each country’s settings, was developed for interviews with journalists, public health advocates, and health policymakers (supplementary file). These guides were designed in alignment with the study’s objectives and were developed by experienced researchers with expertise in media and health research and further reviewed by the mentor and collaborating experts. Two mentors, a senior-level journalist with expertise in health reporting and a university professor of media, mentored the development of the data collection tool.

In Bangladesh, data collection was conducted from 22/10/2024 to 07/01/2025, while in Nepal, data collection took place from 03/09/2024 to 06/03/2025. Interviews were conducted face-to-face, and a few online via Zoom, depending on the participant’s availability and preferences. Interviews lasted approximately an hour in Nepal and between one and one and a half hours in Bangladesh. The interviewers included a media researcher, public health researchers with substantial experience in qualitative research, and a health economist with expertise in urban health. Data was collected until no new information emerged and saturation was achieved.

### Data management and analysis

All interviews were conducted in national languages (Bengali in Bangladesh and Nepali in Nepal) and audio-recorded. The recordings were then transcribed verbatim in their respective languages by trained transcribers, who were oriented and guided by the research team. The transcripts from each country were reviewed against the audio recordings and corrected based on discussions among the research team members. After ensuring accuracy, these transcripts were translated into English. The research team in both countries checked the appropriateness of the translation. The English translations facilitated discussions on analysis plans for both countries. Initially a deductive approach was taken to facilitate a common approach to analysis across the two countries. A priori codebook was developed based on topic guides and refined following a series of discussions among the researchers from both countries. The researchers coded the transcripts and discussed discrepancies as well as new codes that emerged inductively. The research team then reached consensus on a common set of codes, and the draft of the codebook was finalized. The English-translated transcripts were coded using NVivo software (version 14 in Nepal and version 13 in Bangladesh).

Themes and sub-themes were then developed through thematic analysis informed by the six phases outlined by Braun and Clarke [36]. Initially, researchers from both countries familiarized themselves with the data, discussed the common codes emerging from the data, and conducted coding. Upon completion of coding in both countries, the codes were compared between Bangladesh and Nepal to identify similarities and differences. Key messages from each code were discussed within the team to develop initial themes aligned with the objectives. The themes were then discussed, refined, and finalized. Both countries wrote summary analyses based on the discussed themes, and after thorough review and discussion within the team, a cross-country comparative analysis was performed.

### Ethical consideration

This study obtained ethical approval from the Institutional Review Board (IRB) of the BRAC James P Grant School of Public Health, BRAC University (protocol no. IRB-2024-IS-17); the ethical review board of the Nepal Health Research Council (protocol no. 201-2024); and the University of Leeds (MREC 23-057). A participant information sheet was shared with the participants prior to each interview, providing details about the study. Written informed consent was obtained from all participants for the audio recording and note-taking. All ethical procedures were adhered to during data collection and management. Confidentiality and privacy were maintained throughout the study, enabling participants to express their views with confidence and comprehensively.

**Table 1:** Interview summary (n=30)

|  | **Nepal** | **Bangladesh** |
| --- | --- | --- |
| Journalists<br>(n=17) | Print Media (5M, 3F) | Print Media (4; 2M, 2F) |
|  | Online News Portal (2M) | Online News Portal (1M) |
|  | Radio and Television (3M, 2F) | Television (2F) |
|  | Media ownership: 9 private,<br>1 government-owned | Media ownership: 6 private, 1<br>government-owned |
| Policymakers<br>(n=10) | Ministry of Health and Population (2M) | Ministry of Health & Family<br>Welfare (2M) |
|  | Department of Health Services (2M, 1F) | Local Government (2M) |
|  | Academia (1M) | Academia and Planner (1M) |
| Public health<br>advocates (n=3) | None | NGO/CSO (1M) |
|  |  | INGO (1M) |
|  |  | Academia (1M) |
| Total | 15 (10 journalists and 5 policymakers) | 15 (7 journalists, 5 policymakers<br>and 3 public health advocates) |

M=Male; F=Female

### Findings

The findings from Bangladesh and Nepal are organized into four overarching themes. The analysis highlights both shared patterns and context-specific differences in information sources, use, and verification practices; urban health coverage and reporting challenges; and stakeholders’ engagement and the role of journalism in shaping urban health discourse and policy influence.

#### 1. A hierarchy of information access, sources, and verification practices

Across Bangladesh and Nepal, stakeholders including health policymakers, public health advocates, and journalists rely on a range of information sources to inform their understanding of urban health issues. However, the selection and utilization of these sources vary according to professional roles, institutional mandates, and the specific purposes for which information is sought. The majority of the stakeholders mentioned government reports, national surveys, hospital records, and the national health information system—which compiles data from the public healthcare system across primary, secondary, and tertiary levels of care—as the primary formal sources of data and information. In Bangladesh, however, data published by the government institution, although sometimes seen as biased, was considered more useful for policy discussion and advocacy since the government acknowledged it.

> “Because the government doesn’t acknowledge all data and information (evidence), it is easy to discuss the ones the government has acknowledged. It’s also useful for policymaking.” (KII 15, Male, Public Health Advocates, Bangladesh)

In both countries, journalists prioritized government data as a source for information to develop news stories. However, they mentioned that access to government information is increasingly difficult due to bureaucratic hurdles, non-cooperation, and concealment by official sources, necessitating increased efforts to verify information through multiple channels.

> “While the government reported only a few dengue cases, a visit to Dhaka Medical College revealed a much higher number of patients. This highlights the difficulty journalists face in accessing facts when official sources are unreliable, underscoring the need for credible foundations in reporting.” (KII 7, Male, Journalist, Bangladesh)

Furthermore, political influence and control from the government’s institutions restrict journalists’ access to information in both the countries. A journalist in Bangladesh reported that even with permission, news sources may face pressure and potential career consequences, especially when the message contradicts the interests of the organization or the influential people within that particular organization, undermining reporting that emphasizes accuracy and accountability by placing verifiable details over opinion.

> “Even after getting permission, accessing real information was difficult. In one case, when the doctor finally spoke (mentioning the facts), he was soon transferred under pressure from higher authorities.” (KII 4, Female, Journalist, Bangladesh)

Bureaucratic barriers further constrained reporting in both countries. Journalists described repeated visits, delayed permissions, and limited responses from officials, making urban health reporting time-consuming and exhausting.

> “I kept being sent from floor to floor… They said the information couldn’t be given without the director general’s permission.” (KII 10, Male, Journalist, Bangladesh)

When data was insufficient or contested, journalists in both countries also relied on expert opinions and insights.

> “Health information is technical and sensitive, and the available data are often unclear or interpreted differently. In such situations, I consult experts to review my work and confirm my interpretation. Their input is helpful when evidence is insufficient or confusing in some specific cases.” (KII 14, Male, Journalist, Nepal)

These opinions and insights helped journalists develop special and investigative reports as well. However, they emphasized the need to cross-check information from multiple sources for verification to ensure the accuracy and reliability of news.

> “We often don’t rely on only one source. We can rely on the spokesperson of the ministry. However, to verify information, we need to talk to at least 2-3 individuals. Further, documentary evidence is also helpful for information verification.” (KII 2, Female, Journalist, Nepal)

A major challenge across both countries was limited access to reliable, timely, and transparent urban health-related data, which constrained accurate urban health reporting. Bangladeshi journalists, in particular, highlighted data concealment, inconsistencies, and reliance on outdated sources.

> “Journalists face a severe shortage of updated, reliable, and locally relevant urban health data, often relying on outdated research.” (KII 6, Female, Journalist, Bangladesh)

In Nepal, journalists reported that engagement with policymakers was often occasional and constrained by limited access to specialists or experts due to their time unavailability restricting in-depth interaction.

> “At first, it is difficult to approach and meet with health experts who are responsible for providing reliable information.” (KII 1, Male, Journalist, Nepal)

In Bangladesh, peer-reviewed journal publications and development partners’ reports were also often used as sources of evidence by stakeholders. Policymakers used them to understand sector trends and support policy development, although they preferred short, people-centered insights. Public health advocates used these sources to frame issues and identify gaps, and journalists often turned to these sources to authenticate and confirm their data and stories for news development.

> “The articles should be people-centric and written in a simple and understandable language so that we can understand the content. While a clear research approach may be necessary, our primary consideration is whether the article meaningfully engages the public.” (KII 1, Male, Policymaker, Bangladesh)

However, in Nepal, both policymakers and journalists reported limited use of peer-reviewed articles, although they considered them reliable. In both countries, they found it difficult to grasp the technical jargon and experienced challenges in interpreting and using information from research publications.

> “That trend is extremely low; in fact, the practice of using journal articles for information is rare. Personally, I don’t read or take reference of the journal article. Understanding technical language is also difficult.” (KII 9, Male, Journalist, Nepal)

Field visits and primary data collection were widely valued for generating contextual evidence by most stakeholders in both countries. Journalists, in particular, used informal networks and local informants and engaged with affected communities, supported by their long-standing institutional relationships, to access both on- and off-the-record information.

> “During the anti-discrimination student movement in July 2024, there was a lot of secrecy around post-mortems (reports for the deaths). Some deaths were not officially listed, which made cross-checking difficult. Verifying information required building relationships with mortuary staff or an autopsy technician.” (KII 4, Female, Journalist, Bangladesh)

In both contexts, health policymakers and public health advocates reported news media as an important source of health information but largely viewed it as a starting point rather than definitive evidence, due to concerns about the limited number of well-trained, health-focused journalists and the risk of misrepresentation or selective reporting.

> “The journalists having experience in the health sector are insufficient, which leads to the possibility of misinterpretation of information/evidence and research findings.” (KII 7, Male, Health, Nepal)

However, when evaluating information and evidence, policymakers and public health advocates reported placing greater trust in well-established health-beat journalists and newspapers, often assessing credibility based on the reputation of the outlet and the strength of its editorial board.

> “They edit both the language and the information. In one of my articles, an incorrect global population figure was corrected during editing. Because such newspapers have a good editorial board and publish content after proper review; their information is reliable.” (KII 15, Male, Public Health Advocate, Bangladesh)

Most stakeholders also noted that social media platforms, such as YouTube, WhatsApp Groups, and online news articles, are more popular due to their ease of use. Digital media appeared handy. However, concerns about source credibility remained.

> “Social media, along with online media, is the recent trend. But to what extent should we believe them? I do not think online media can be widely trusted, as there are already established news media such as television and newspapers, where information is more reliable.” (KII 8, Policymakers, Female, Nepal)

#### 2. Insufficient urban health coverage and reporting challenges

Journalists in both Bangladesh and Nepal mentioned urban health reporting as important yet consistently under-prioritized within the media landscape. It was rarely treated as a primary reporting area within the health beat. In Bangladesh, although health journalism has existed for long but gained greater visibility following the COVID-19 pandemic, coverage of urban health issues remained fragmented and largely event-driven rather than issue-focused and follow-up story-driven.

> “As such, urban health is typically ranked lower in priority (in reporting), perhaps around seventh or eighth. Issues such as recruitment, transfers, and corruption dominate the discourse in Bangladeshi health content.” (KII 5, Female, Journalist, Bangladesh)

Similarly, journalists in Nepal mentioned that urban health received limited and inconsistent attention, with inadequate in-depth exploration.

> “I don’t feel like people have really worked on exploring in depth in the area of urban health.” (KII 1, Journalist, Male, Nepal)

Additionally, they highlighted that urban health is not considered a distinguished area within the health beat. Reporters were often required to cover a variety of topics, limiting continuity in urban health reporting.

> “There is very little reporting when a beat focuses specifically on health as a distinct social issue. There is no dedicated urban health beat where reporters consistently focus on just this area. If someone reports on it today, they might move on to another topic tomorrow.” (KII3, Journalist, Male, Nepal)

Despite the low prioritization, certain media outlets in Bangladesh reported relatively regular coverage of urban health-related issues. Some private television channels also introduced dedicated health programs featuring various doctors, titled “Prescription.” Additionally, one media outlet initiated a hospital visit program that offered guided visits to hospital wards to provide insights into healthcare delivery, referred to as a “Ward Tour.”

> “There were also supplements, like lifestyle… or dedicated health pages where many types of information were published, and that’s where I mainly worked. For example, a leading daily newspaper features a dedicated health page published weekly, covering topics such as healthy living, doctor–patient question-and-answer sessions, and related health information.” (KII 2, Male, Journalist, Bangladesh)

In both Bangladesh and Nepal, urban health reporting was largely dominated by outbreaks and hospital-based issues. Commonly covered topics included infectious diseases (e.g., dengue, COVID-19, diarrhea, and cholera), air pollution, waste management, health facility problems, reproductive and maternity care, health financing, and access to health services.

> “Urban health spans illnesses, viral-borne diseases, pollution, hygiene, and nutrition, alongside challenges such as inadequate sanitation, poor hygiene maintenance, and dense slum settlements in cities like Dhaka. It also includes the health needs of diverse populations, including children, women, the elderly, and the floating population.” (KII 05, Female, Journalist, Bangladesh)

However, journalists in both countries acknowledged that issues related to non-communicable diseases (NCDs), substance abuse, and chronic illnesses receive less attention, making it difficult to generate media interest despite their long-term health and social impact.

> “Often the issues of covering NCDs, substance abuse, and cancer are neglected as they are not expressed in great numbers, although their impact is long-lasting.” (KII 14, Journalist, Male, Nepal)

Journalists in both countries also noted that, even amid news of competing priorities, they worked to incorporate the voices of marginalized and underprivileged urban populations, such as slum dwellers and the urban poor.

> “For instance, a certain percentage of the population faces significant health risks, and I ensure that this is presented (in my reports). They consistently receive the highest priority in my reporting.” (KII 12, Female, Journalist, Bangladesh)

While in Nepal, some journalists have reported that urban health concerns are often overshadowed by the assumption that city residents are relatively affluent and health literate, overlooking the issues of urban slums and poor populations. As one journalist expressed:

> “There is a misconception that everyone living in the city is wealthy and health literate, resulting in urban health issues related to slum or poor populations being less reported.” (KII12, Female, Journalist, Nepal)

In Bangladesh, journalists reported direct pressure from powerful actors, including intimidation, denial of access, and personal threats, particularly when reporting on sensitive or stigmatized urban health and social issues. Such threats were reported less frequently by Nepali journalists.

> “When we work on issues like homosexuality or child marriage, people are scared to speak, the data isn’t accurate, and threats become part of our work—we report by accepting that risk.” (KII 12, Journalist, Bangladesh)

Most journalists interviewed described broader structural challenges, including limited institutional support for fellowships, lack of training opportunities in health journalism, and professional development, which restricted journalists’ skill enhancement and quality reporting.

> “Unlike in other areas, there are very limited professional fellowship opportunities to support journalists in health reporting. Urban health issues could have received better attention if such fellowships were provided.” (KII 1, Male, Journalist, Nepal)

Additionally, in both countries, journalists highlighted financial constraints, including low salaries and a lack of additional remuneration, as key barriers to investigative journalism or specialized urban health reporting.

> “If we keep urban health issues as our priority, then (additional) workforce is required to prioritize and push these issues forward, which is lacking in the current situation due to underpayment.” (KII 12, Female, Journalist, Nepal)

Moreover, health news coverage also depended on consistent funding and adequate financial backing to ensure its continuity. As one journalist mentioned,

> “It may appear that newspapers prioritize health, but in reality, this happens because of funding. Health coverage continues where there is strong subsidy or advertisement support.” (KII 2, Male, Journalist, Bangladesh)

However, in both countries, special reports give journalists recognition and a sense of identity, which still motivates them to continue pursuing investigative reporting, unlike routine daily news without bylines.

> “The greatest benefit of special reporting is the recognition these stories give to me because these are published under my byline defining my professional identity. Routine daily reports, by contrast, are typically anonymous.” (KII 07, male, Journalist, Bangladesh)

While a few journalists in Nepal noted the challenge of negative public perceptions and declining prestige and respect, which have undermined journalists’ professional pride, leaving even committed journalists feeling undervalued.

> “Because of some negative people who do not engage in quality reporting, even professional journalists sometimes cannot feel proud of their profession. The pressure, prestige, and respect that journalism had in the past are no longer the same.” (KII 1, Male, Journalist, Nepal)

#### 3. Media-stakeholder engagement practices

In both Bangladesh and Nepal, policymakers, public health advocates, and journalists interacted and engaged along functionally different pathways. Such interactions were shaped by perceptions of stakeholder expertise and roles, as well as the credibility and use of information in policy decisions.

In Bangladesh, health policymakers reported systematic and institutionalized engagement with researchers, technical experts, and advocacy organizations, particularly for evidence generation, technical analysis, and policy shaping. Journalists, by contrast, were primarily engaged for information dissemination and public communication and awareness, rather than policy deliberations. Although senior and more experienced reporters were occasionally consulted, junior journalists generally perceived their role as largely that of an observer.

> “When we conduct research, such as on National Health Accounts, we highlight out-of-pocket expenditure and catastrophic health expenses. During this dissemination, they [journalists] engage. But for our other internal research, we don’t involve them.” (KII 9, Male, Policymaker, Bangladesh)

Health policymakers across both contexts also expressed concerns about inconsistent journalistic practices, noting that while some media outlets coordinated closely, others published without adequate verification with the concerned organizations.

> “The major media outlets that produce good-quality news coordinate with us. However, some media organizations publish information without proper research, proper data analysis, or thorough verification of information.” (KII 4, Female, Policymaker, Nepal)

In contrast, journalists mentioned initiating regular contacts with policymakers and public health advocates to incorporate their opinions, which they believed was required for the development of an acceptable report.

> “Yes, regularly, and in special reports, each of them provides comments; their opinions are taken, and they must be consulted. Without this, your report won’t be accepted.” (KII 7, Male, Journalist, Bangladesh)

However, in both countries, stakeholder engagement was also relationship-driven, and the journalists could also gain access to health experts and officials based on their connections.

> “I even contacted the director directly to confirm the information, and a professional connection facilitated access.” (KII 4, Female, Journalist, Nepal)

In both countries, interaction and engagement of journalists, policymakers, and public health advocates occurred through specific platforms and occasions, rather than through formalized institutional collaboration with media.

In Bangladesh, journalists were commonly engaged through formal events such as national observance days (e.g., Universal Health Coverage Day, World Tuberculosis Day), research dissemination events, press briefings, and health campaigns related to dengue, COVID-19, or emerging public health threats. Policymakers described journalists’ presence in these spaces as essential for visibility and outreach, though their role remained largely observational.

> “On Universal Health Coverage Day on December 12, we invite journalists for a major event. At the same time, journalists also approach us.” (KII 9, Male, Policymaker, Bangladesh)

Similarly, in Nepal, policymakers reported mobilizing media during vaccination campaigns, new health program launches, and public awareness programs, recognizing journalists as critical intermediaries between the health system and the public.

> “Almost every major or mainstream media outlet reports on public health events. We need media support when formulating or endorsing policy, and we even mobilize them ourselves.” (KII 5, Male, Policymaker, Nepal)

Across both contexts, crises and urgent public health threats, such as disease outbreaks, water shortages, or service disruptions, emerged as key moments of increased interaction. Journalists actively sought expert opinions, while advocacy actors and policymakers responded to media inquiries to manage public concern.

> “If a waterborne infectious disease is spreading… journalists call us, seek information from us, or take telephone interviews to seek our advice on how to address the situation.” (KII 3, Male, Advocacy Actor, Bangladesh)

#### 4. Role of media in influencing urban health policy decisions

Most journalists strongly believed that the news media played an important role in bringing neglected or critical health issues into the spotlight, drawing public attention and placing pressure on government actors to respond in both countries. Despite limited formal inclusion in decision-making, stakeholders acknowledged that media exert indirect but significant influence on urban health agendas by shaping public discourse and generating pressure for action.

In Bangladesh, stakeholders cited multiple examples where sustained media coverage led to concrete policy or service delivery responses by raising awareness and serving as a watchdog, particularly during crises. Media coverage during COVID-19 that drew attention to gaps in intensive care unit (ICU) capacity, oxygen availability, and protective equipment was widely seen as instrumental in accelerating government action and resource allocation.

> “Every public hospital has increased ICU beds compared to before and after COVID. ICU beds have been increased. Oxygen plants have also been significantly expanded. These full-fledged improvements happened solely because of news reporting.” (KII 14, Male, Policymaker, Bangladesh)

Stakeholders perceived that media reporting also influenced urban health planning, vaccine introduction, and accountability mechanisms, including dengue surveillance, mosquito control monitoring, and service provision in marginalized communities in both countries.

In the similar way, journalists in Nepal described their role as bringing neglected issues into public view, thereby compelling government attention.

> “After I wrote the news about pesticides in vegetables, the District Agriculture Office immediately provided training to the farmers, and monitoring started in the market.” (KII 4, Female, Journalist, Nepal)

Policymakers acknowledged the media as a “pressure-raising group” and agreed that media coverage played a role in raising awareness and creating public pressure, which influences decision-making, particularly in urgent health matters. They shared examples where the media had a positive impact, such as agricultural inspections following reports on pesticide use, infrastructure responses after emergency health reporting, the introduction of the Smoking and Tobacco Control Act, and the ‘No Horn Policy’ in Nepal.

> “News media plays a vital role in health policy decisions because people’s perceptions depend on how media gives coverage. If the media writes about prevailing issues, the public becomes conscious and decides cautiously. It does create an impact. It also creates pressure, highlighting that this work is happening in this field.” (KII 6, Male, Policymaker, Nepal)

Most policymakers agreed that the news media influences policy decisions; however, one of the policymakers in Nepal shared the view that direct policy change based solely on media reporting remains limited.

> “The case for policy change is very low, just because it was written in the news. The media facilitates both bringing and not bringing a certain policy.” (KII 7, Male, Policymaker, Nepal)

Stakeholders in both countries highlighted a lack of meaningful collaboration, making it difficult to observe direct policy impact. In Bangladesh, policymakers noted a perceived decline in the media’s influence over time, attributing this — though still debated — to political dynamics, bureaucratic inertia, and reduced electoral consequences of media exposure.

> “Twenty years ago, a newspaper report could trigger immediate action. However, at present, media reports are no longer associated with electoral voting influence, so the government shows little concern.” (KII 8, Male, Policymaker, Bangladesh)

In Nepal, mistrust between journalists and policymakers and complaints of policymakers regarding unethical journalistic practices sometimes weaken relationships and collaboration, particularly at provincial and local levels.

> “The media are liars as well… they take benefits… Not everyone does it in the right way.” (KII 6, Male, Policymakers, Nepal)

Despite these challenges, both settings showed emerging spaces for more inclusive engagement. In Bangladesh, multi-stakeholder platforms and advocacy coalitions increasingly brought journalists into policy dialogues as active contributors rather than passive disseminators.

> “I participated as a media representative, and interestingly, two of the recommendations I had previously shared were discussed during the meeting. Such invitations were extended to several of us from the media.” (KII 4, Female, Journalist, Bangladesh)

However, the journalists in both countries still expressed a desire for more structured and meaningful engagement—particularly during the early stages of policy formulation, rather than being briefed only after decisions were made.

> “Journalists are rarely invited to attend ministry meetings (only on limited occasions). Instead, they receive a briefing afterwards, where decisions and discussion points are summarized. However, if I were able to attend the entire meeting, I could better understand the full scope of the discussions, the tone of the deliberations, and the challenges being raised across different areas.” (KII 7, Male, Journalist, Bangladesh)

## Discussion

The role of media and research in shaping urban health policy reforms, especially in knowledge dissemination and stakeholder communication, has long been acknowledged [13, 14]. However, limited evidence exists on how news media access, interpret, and use evidence as well as how they engage with stakeholders to inform urban health policies, particularly in low- and middle-income countries (LMICs) [14, 37]. Our study contributes to addressing this gap by examining the challenges related to data accessibility, evidence use, and stakeholder engagement among policymakers, public health advocates, and journalists in Bangladesh and Nepal. Based on interviews with stakeholders, findings suggest that the media function as a crucial intermediary between evidence and policy processes, but they work within structural constraints, credibility issues, and episodic rather than institutionalized engagement. Moreover, the cross-country comparison highlights common journalistic practices and context-specific dynamics that shape urban health reporting and determine whether—and to what extent—it influences urban health policy decisions.

In both countries, four interconnected dynamics became evident: first, uneven access to information and fact-checking verification practices; second, the insufficient prioritization of health and urban health reporting; third, limited institutionalized engagement among the stakeholders; and fourth, the indirect influence of news media on urban health policy decisions and

### Uneven access to evidence and fact-checking practices

Stakeholders in both Bangladesh and Nepal relied on diverse sources with varying levels of quality, reliability, and accessibility. They also use government reports and survey datasets of respective government agencies. Besides, journalists also use field visits and informal networks and experts’ opinions for fact-checking. Similar to other low- and middle-income countries [38, 39], urban health reporting is often pulled from data generated by government agencies, considered a key source of evidence, and generally perceived as more acceptable because of government acknowledgement despite room for bias [38, 39]. However, journalists in both countries reported persistent challenges in accessing these data, often due to bureaucratic hurdles and political sensitivities, which eventually increased the gaps between evidence and media narratives. Similar patterns have been observed in global health journalism, where journalists struggled to access health data while navigating power structures [28, 40, 41].

Confronted with these challenges, journalists in both countries mentioned consulting multiple sources to cross-verify information using a triangulation approach. These practices align with international journal standards that emphasize cross-verification in contested or data-scarce environments [42–44]. Nevertheless, despite journalists’ efforts to verify information, public health advocates and policymakers in both countries expressed concerns about the credibility of some media coverage, which is sometimes not backed by reliable information.

Stakeholders further mentioned how editorial gatekeeping, political pressure, and threats influenced media reporting, especially in Bangladesh. This was consistent with other regional studies in Bangladesh about press freedom and political influence [45–48]. In Nepal, journalists reported fewer direct threats but highlighted weaker specialization in health journalism and growing concerns about/over credibility. However, previous studies in Nepal showed that, while credibility issues persisted [49, 50], journalists were also subject to threats and assaults [51]. These contextual variations demonstrate that news reporting practice and policy reforms are shaped by political culture, institutional practices, and power relations.

Journalists and other stakeholders in Bangladesh also referred to using peer-reviewed research articles as information sources, but the practice was particularly limited in Nepal. As reported by the journalists, technical jargon and complex language of the journal articles were main reasons for limited use. Moreover, policymakers wanted research to be practical and people-centric and believed evidence uptake is higher only when evidence (research findings) is shared in a timely manner. These findings align with global literature indicating that, for non-academic audiences, the relevance, clarity, and timely accessibility of research output are important, as well as how the narratives are framed and presented [52–54].

### Insufficient prioritization of urban health in media reporting

Despite the growing burden of urban health challenges, participants in this study perceived urban health reporting as under-prioritized in both countries. Media coverage was narrow and focused on visible issues such as disease outbreaks, hospital crises, pollution, and sanitation concerns. Urban health issues, including chronic conditions such as NCDs; mental health problems; substance use; waterlogging and related health concerns; the vulnerabilities of marginalized populations in the urban context; and other social determinants, receive little attention. This pattern is consistent with broader trends in global health journalism, where news coverage tends to focus on sudden events and serious situations such as disease outbreaks or health system crises, because these issues quickly capture public and media attention. In contrast, slower and less visible health challenges, including chronic diseases and persistent gaps in healthcare services, often receive comparatively less media attention [55–57].

Further, journalists in Nepal also noted a misconception that urban residents are relatively well-off, which leads to less emphasis on intra-urban inequities and limited prioritization of urban health issues in reporting practices. Although journalists in Bangladesh did not specifically raise this issue, evidence from urban health studies suggests that aggregate/overall urban indicators often overlook disparities within cities [58–60]. Poor urban populations often face worse health outcomes than rural residents due to exclusion, informality, poor infrastructure and living conditions, and fragmented services [61]. These issues receive limited news coverage unless crises occur [58, 62–65]. Our study thus highlights the need for more equity-focused framing in urban health reporting to bring such disparities in limelight beyond crisis moments.

In our study, journalists also reported challenges related to low salaries, limited fellowship opportunities, weak institutional support and training, and dependence on advertising or donor funding. This reflects broader patterns in LMIC contexts, where urban health journalism remains under-resourced and financial and institutional constraints contribute to the under-prioritization of urban health reporting [66]. These structural barriers restrict the development of specialized or investigative urban health reporting, often essential for health policy reforms.

Despite these constraints, some media outlets in Bangladesh and Nepal have introduced health as a separate beat, including newspaper health pages and special episodes of television programs. However, ‘urban health’ is not commonly recognized as a distinct reporting area within newsroom structures. As a result, the diverse and often complex health challenges faced by urban populations, especially the marginalized groups, may receive limited and inconsistent coverage. This pattern mirrors that of other South Asian countries and LMICs, where health reporting, particularly urban health, is often not consistently prioritized. Even where health-focused reporting arrangements exist, they often lack adequate resources and well-trained specialized health reporters to ensure sustained, investigative, and in-depth coverage of health issues [8, 17, 67–69].

### Limited institutionalized engagement among the stakeholders

Policymakers and public health advocates primarily engaged journalists for dissemination purposes and public communication, while journalists sought their help for collecting evidence, cross-verifying information, and obtaining legitimacy. This limited and occasional engagement reduced the possibility of evidence-based discussion before policies were finalized. However, in Bangladesh and many other developing countries, multi-stakeholder platforms and advocacy coalitions are attempting to engage media as active participants in policy discussions, rather than as passive disseminators [70, 71]. While the media in both countries have contributed to raising awareness and shaping public health discourse, structural barriers—including power dynamics, credibility issues, and limited institutional engagement—continue to constrain their ability to contribute to urban health policy reforms. Strengthening more structured and continuous institutionalized engagement among the stakeholders, especially among the media and health policymakers, could help address these limitations. In addition, improving mechanisms for transparent communication and cross-verification of information can enhance trust among the stakeholders. Evidence suggests that transparent communication contributes to greater public trust and policy legitimacy [66–68]. Such efforts are especially important in rapidly urbanizing LMIC settings, where health inequities are widening, and evidence-informed policy is essential for achieving equitable urban development [2, 32, 39].

Creating an enabling environment for independent and well-resourced journalism is also critical. Media operating free from political pressures and power plays are better positioned to investigate, frame, and communicate evidence to inform urban health reforms [28, 71]. In this context, investments in dedicated urban health reporting, along with training and fellowship opportunities for journalists, could strengthen their skills and capacity to cover urban health issues beyond episodic events and crises [8, 68, 72].

### The Indirect Influence of News Media on Urban Health Policy Decisions

The media’s role in influencing health policy was generally described as indirect but meaningful. Stakeholders in both countries highlighted instances where media attention generated public pressure and accelerated government response, particularly during health emergencies. In LMIC health systems, the media have shaped public health discourse, played the role of watchdog, and exerted pressure for generating actions in different circumstances [13, 32].

However, there’s little evidence that the media meaningfully contributes to the urban health policy process in LMICs beyond information dissemination, owing to concerns over credibility, mistrust, and poor communication among stakeholders as seen in both Bangladesh and Nepal [13, 73, 74]. As a result, media influence may be both direct and indirect, but more often it is indirect rather than direct and proactive. Consequently, the influence of the media often becomes most visible during crises, when rising public attention pushes policymakers to act.

### Strength and limitation

A strength of this study is the cross-country comparative design, along with the inclusion of multiple stakeholders (health policymakers, public health advocates, and journalists) and triangulating the findings, which provided rich insights into the media–policy interface in two South Asian LMIC contexts. These insights will support the research partner organizations involved in the study and other institutions and think tanks to effectively engage with both media and policymakers in the region, promoting evidence-informed decision-making and strengthening urban health systems. Additionally, by generating this evidence, the study adds to the broader global health journalism and urban health literature.

This study has some limitations. First, this study did not capture the perspectives of the general population or elected officials, whose views and decisions are often shaped by media-influenced public opinion. Their absence limits a more comprehensive understanding of the broader media–policy ecosystem. Second, responses may also reflect social desirability bias, particularly in discussions of political pressure or credibility from the journalists’ and policymakers’ perspectives. Third, although the cross-country comparison highlights common patterns and context-specific differences, the findings may not extend to contexts outside Bangladesh and Nepal. Finally, some interviews were conducted online due to participant time availability and preferences, which may have influenced the depth of information compared to face-to-face interviews.

### Conclusion

This study highlights that health reporting in Bangladesh and Nepal relied on diverse information sources but faced persistent challenges, including limited access to reliable data, credibility concerns, and technical and skills-related challenges. Urban health reporting is inadequately prioritized, as the coverage mainly revolves around visible issues (e.g., outbreaks, disasters, etc.) and crises rather than chronic health conditions or long-term urban health inequities and challenges. Engagement between journalists and other stakeholders also tended to be event-driven and episodic, with journalists primarily working as intermediaries for public awareness rather than active participants in the policy formulation process. While news media influence on urban health policy decisions was evident, it remained somewhat limited by structural barriers, political dynamics, and weak institutional collaboration.

This underscores the need for prior and more systematic engagement of journalists in the policymaking process to support evidence-based urban health policy decisions. Further, improving transparency and access to reliable government data could strengthen this process. Researchers should also share findings with journalists in more accessible ways and formats such as policy briefs, media summaries, and community-oriented stories to ensure they inform public debate in a timely and relevant manner. Further, journalism should remain independent and free from political influence, with health or urban health issues receiving higher priority in news coverage. Further acknowledgement of responsible health reporting by policymakers and public health advocates may further motivate the media to contribute more meaningfully to public debate and policy reform processes.

## Data Availability

Data availability statement: The data generated and analyzed from IDIs and KIIs in this study are not publicly available because they contain information from the participants who consented to the use of the data for this study and thus would not allow for public storage. Data will be available from the corresponding authors on request. Data are stored at the institution’s database, and upon reasonable request, it can be accessed upon request and procedure. Persistent, long-term storage and availability of the data will be ensured within the organization’s secure database system of both the institutions engaged in this research.

## Acknowledgments

We sincerely thank all the journalists, policymakers, and public health advocates in Bangladesh and Nepal who participated in this study and shared their time and experiences. We also acknowledge the current and former team members at BRAC James P Grant School of Public Health, BRAC University, HERD International, and ARK Foundation whose contributions supported different stages of this study. We thank Shishir Moral from Bangladesh and Sudhamshu Dahal from Nepal for their mentorship and valuable guidance throughout this study.

